# Rhythms of Brain Impedance

**DOI:** 10.1101/2022.10.16.22280780

**Authors:** Filip Mivalt, Vaclav Kremen, Vladimir Sladky, Jie Cui, Nicholas Gregg, Irena Balzekas, Victoria Marks, Erik K. St Louis, Paul E. Croarkin, Brian Nils Lundstrom, Noelle Nelson, Dora Hermes, Steven Messina, Samuel Worrell, Thomas J. Richner, Benjamin H. Brinkmann, Timothy Denison, Kai J. Miller, Jamie Van Gompel, Matt Stead, Gregory A. Worrell

## Abstract

Brain impedance is a fundamental electrical property that depends on tissue extracellular volume. We tracked impedance, behavioral state, and epileptiform activity in humans using an investigational device and identified behavioral state dependent impedance oscillations spanning hours to weeks in amygdala, hippocampus, and thalamus. Impedance reaches a minimum in slow wave sleep, is intermediate in rapid-eye-movement sleep and maximal during wakefulness consistent with previously observed extracellular volume changes in rodent glymphatic system.

## MAIN

Impedance is an extrinsic tissue property important for characterizing the resistance to endogenously^1^ and exogenously^2^ generated ionic currents in the brain. The impedance influences the spatiotemporal dynamics of extracellular ionic currents giving rise to local field potentials^1^ and the volume of tissue activated by electrical brain stimulation (EBS)^2^. The voltage generated in response to an applied current can be used to measure brain impedance, and at the scale of clinical EBS electrodes (∼mm^2^) depends on the electrode-tissue interface and ionic current conductance through the intracellular, vascular and extracellular space (ECS) compartments of brain tissue^3^. The impedance of rodent^3,4^, non-human primate^1^ and human^5^ brain tissue over the frequency range of physiological local field potentials is primarily resistive^1,6^ and largely determined by the ECS volume fraction^4,7^. The interstitial conductive-fluid filled ECS creates an electrical network that is increasingly recognized for its role in the brain glymphatic system and sleep-dependent brain health, such as clearing potentially neurotoxic metabolites that accumulate during wakefulness and pathologic proteins associated with neurodegenerative disorders.^8–11^

The temporal dynamics of human brain impedance (*Z*) have received relatively little attention despite being an integral factor in brain electrophysiology. Prior human studies report higher tissue conductance in brain regions generating seizures^5^ and focus on the role of impedance in determining the volume of tissue activated by EBS^2^. In patients undergoing therapeutic EBS the impedance was reported to increase over a few weeks after electrode implantation to a constant stable value^12^. These studies in humans, however, are limited to sparsely sampled measurements and are inadequate for systematic investigation of impedance temporal dynamics.

Two-photon imaging in rodent cortex shows the ECS volume increases by ∼60% in the transition from wakefulness to slow wave non-rapid eye movement (NREM) sleep^8^. Similar investigations are not possible in humans. Increased ECS volume in NREM compared to REM sleep was also proposed as the mechanism underlying electrical impedance increases observed during NREM to REM transitions in rodent brain ^3,4^. We hypothesized that if behavioral state transitions drive ECS volume changes in the human brain glymphatic system they would manifest as impedance *Z* cycles with lower values in NREM sleep compared to wakefulness and REM sleep. To investigate this hypothesis we continuously tracked impedance *Z*, behavioral state^13^, and epileptiform activity^14^ in multiple limbic brain structures (anterior nucleus of thalamus (ANT), amygdala (AMG), and hippocampus (HPC)) over multiple months using an investigational implantable neural sensing and stimulation device in five people with drug resistant focal epilepsy living in their natural home environments (Fig. 1 and supplementary figs. 1 - 4).^14–16^

**Figure 1:**
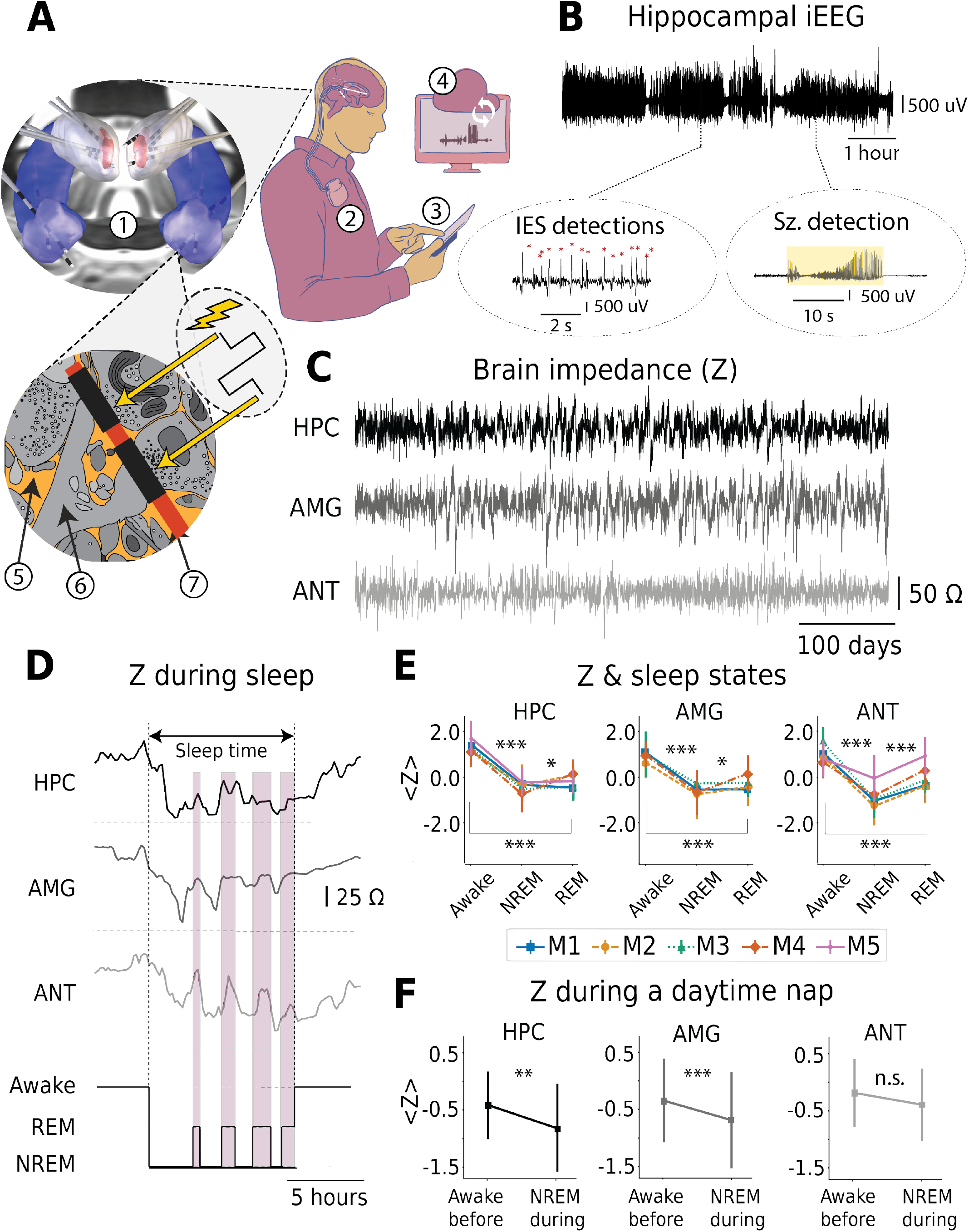
Behavioral State Dependent Brain Impedance in Humans. Brain impedance is decreased in NREM slow-wave sleep compared to REM sleep and wakefulness in limbic brain regions. **A)** Schematic of brain co-processor using investigational INSS device (Medtronic RC+S™ Summit) integrated with local (tablet and smartphone) and distributed cloud computing infrastructure. The system consists of (1) electrodes implanted in bilateral AMG, HPC, and ANT. (2) INSS device implanted in a surgically created sub-clavicular pocket. (3) Custom software application (epilepsy patient assistant) running on a smartphone provides local computing and bi-directional connectivity between the INSS, patient, and (4) cloud data and analytics platform. *Inset:* Tissue impedance, Z = *V/I* is determined from the voltage response (*V*) to an injected current (*I*), and depends on brain microenvironment (5) extracellular, (6) cellular and vascular compartments) surrounding sensing and stimulation electrode (7) (not to scale) **B)** Representative sample of intracranial LFP from HPC. The insets (circled) show automated IES and seizure detections. **C)** Representative HPC, AMG and ANT impedance time series (z-score with 2-hour median filter) from subject M1 living in home environment **D)** Hypnogram from automated behavioral state classifier trained, validated, and tested using human expert visual scoring of simultaneous intracranial LFP and polysomnography. Representative example of HPC, AMG and ANT impedance over one night with four cycles of NREM and REM. The impedance is reduced in NREM sleep compared to REM and wakefulness in AMG, ANT and HPC. **E)** Comparison of behavioral state-dependent z-score impedance(*<Z>*) in NREM, REM and Wakefulness. The AMG, ANT and HPC impedance is lowest in NREM, intermediate in REM and highest in wakefulness (p<0.05, ANOVA and two-way Mann-Whitney test with Bonferroni correction, see supplementary figure 7 for individual subjects) **F)** Four of five subjects took daytime naps that were used for analyzing the effect of behavioral state on impedance. Similar to nighttime NREM sleep the impedance is decreased in NREM sleep during naps compared to wakefulness before napping in the AMG and HPC. (*p < 0.05, **p < 0.01, ***p < 0.001). **Abbreviations:** Amygdala (AMG), Anterior Nucleus of Thalamus (ANT), Brain impedance *Z*, Hippocampus (HPC). Implantable Neural Sensing and Stimulator (INSS), Local field potential (LFP). Non-rapid eye movement (NREM) and rapid eye movement (REM) sleep. Interictal epileptiform spikes (IES), Ohms (Ω), and hour (hr.), Z-score of impedance (<Z>).

We sampled bipolar and monopolar two-point impedance^4^ in AMG, ANT and HPC in five people with focal epilepsy by measuring the voltage response to a charge balanced square-wave current pulse (0.4mA, 80μsec pulse width) delivered every 5 - 15 minutes over more than 79 months (15.39 ± 9.81 months) (Fig. 1A&C and supplementary fig. 3). We excluded electrode contacts used for therapeutic EBS, involved in seizure onset, with high impedance (>5,000 Ω), or frequent artifacts, leaving 45.0% (36/80) of the electrode contacts (5 AMG, 14 HPC and 17 ANT) across 5 subjects for analysis (Supplementary fig. 2). To avoid impedance changes related to surgical implant effect^10^ or seizure activity^17^ (see supplementary figs. 4 - 6) we exclude data within 15 days of device implant and 24 hours before or after seizure activity, leaving 1,382 continuous 24-hr epochs (33,168 hours) from 5 subjects (276 ± 215 24-hr epochs) to investigate ultradian, circadian and infradian impedance cycles.

We previously demonstrated accurate behavioral state classification (wakefulness, REM sleep, NREM slow-wave sleep, or indeterminant) with a Naïve Bayes behavioral state classifier developed using simultaneous intracranial LFP recordings and polysomnography^13^. The impedance during wakefulness, REM and NREM sleep was investigated during the initial one month baseline period without EBS (Fig. 1D, E & F). The AMG, ANT and HPC impedance is lowest in NREM, intermediate in REM and highest in wakefulness (Figure 1E). The impedance was lower in NREM sleep compared to wakefulness (−19.09 ± 13.52 Ω, p<0.001) and REM sleep (−13.78 ± 13.01 Ω, p<0.001) (Supplementary fig. 7). Four subjects took afternoon naps and the automated behavioral state classifier identified daytime nap NREM slow wave sleep episodes (nap onset 2:06 PM ± 1.2 hr.). The impedance in the subjects taking daytime naps was decreased in NREM slow wave sleep compared to wakefulness before naps in HPC, and AMG (Fig. 1F).

Spectral analysis of AMG, ANT and HPC impedance timeseries show oscillations with ultradian (3.47 ± 1.40 hr.), circadian (24.01 ± 0.39 hr.) and infradian (13.09 ± 8.14 days) periodicities (Figure 2A, B and supplementary figs. 8 & 9). The circadian oscillation is regular with little variation in the cycle period over the duration of recording (Fig. 2A, middle panel). The average impedance (30 min. sliding window with 10 min overlap) increases over the course of the day to a maximum at 7:04 PM ± 3:55 hr. in the evening and decreases overnight, reaching an early morning minimum prior to waking at 6:15 AM ± 1:56 hr. (Fig. 2B and supplementary fig. 8). The average impedance difference between the daytime maximum and night-time minimum was 43.06 ± 16.72 Ω, 46.43 ± 15.72 Ω, and 48.94 ± 16.43 Ω for AMG, HPC and ANT, respectively and corresponds to 8.32 ± 1.45 % change in average impedance (AMG: 9.01 %, HPC: 9.64 %, ANT: 6.30 %) (Supplementary fig. 8).

**Figure 2.**
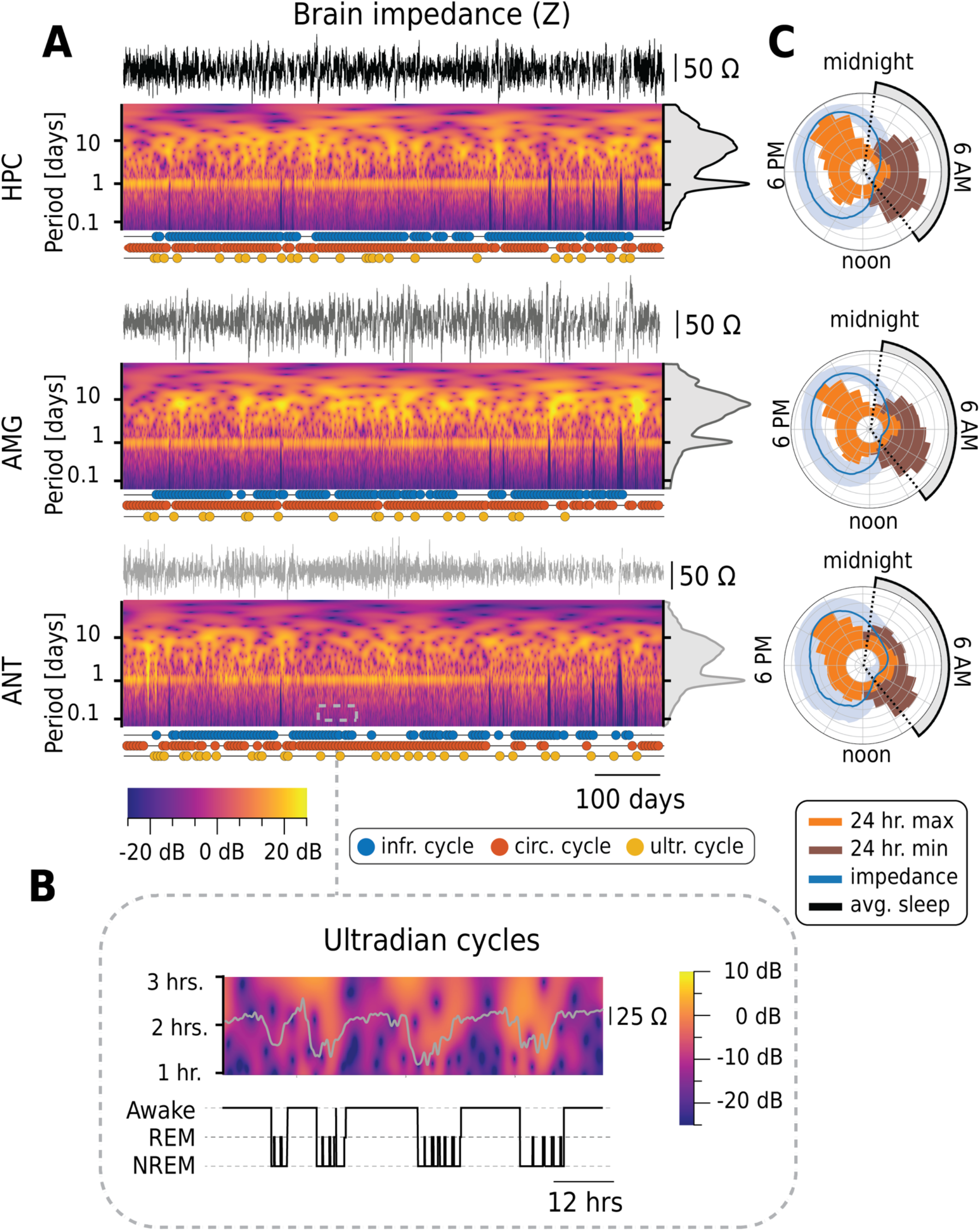
Multiscale Rhythms of Human Brain Impedance. There are ultradian (< 24 hrs.), circadian (∼24 hr. cycle period) and infradian (>24 hr. cycle period) limbic brain impedance oscillations. **A)** Three panels for HPC, AMG and ANT impedance: *Top:* Impedance time series. *Middle:* Continuous wavelet transform (CWT) of impedance time series demonstrating multiscale oscillations with the circadian period oscillation most prominent and stable. Middle-right: Amplitude index (gray) showing the strong circadian peak and broad infradian band. The CWT applied to multiple month impedance records does not capture the ultradian activity well (see Fig. 2C). *Bottom:* Significance test using F-score for ultradian (yellow dots), circadian (red dots), and infradian (blue dots) oscillations (Supplementary figure 9). Consecutive window of data (5 days for ultradian & circadian and 100 days for infradian) are tested for evidence of oscillations. There is a strong, persistent circadian pattern in the limbic network impedance. Ultradian and infradian rhythms are present, but more variable over the multi-month recording. **B)** Blow-up of CWT over 3 days showing a strong ultradian oscillation (∼ 1.25 - 3 hrs.) not visually evident in the long-term amplitude index and CWT. **C)** Polar impedance *Z* plots (blue) and maximum (orange) and minimum (brown) value histogram shows 24-hr pattern with *Z* increasing during the day and decreasing at night during sleep (black). *Z* reaches maximum values in late afternoon or early evening and minimum values during morning prior to wakefulness in the AMG, HPC and ANT for all subjects. **Abbreviations:** Brain impedance *Z*. Amygdala (AMG), Anterior nucleus of thalamus (ANT). Continuous Wavelet Transform (CWT), Hippocampus (HPC), rapid eye movement (REM) sleep, non-rapid eye movement (NREM) sleep. Ohms (Ω). Hour (hr.).

The ultradian and infradian impedance cycles have more variable cycle periods over the multiple month recording (Fig 2B, C and supplementary fig. 9). The ultradian cycles are poorly visualized in the average spectral power density or continuous wavelet transform calculated from the long-term impedance time series (Fig. 2A, middle panel; right, amplitude index(gray)) reflecting the variability of the ultradian cycle periods and durations. Ultradian oscillation are, however, clearly evident in shorter time window analysis spanning multiple days (Figs. 1D & 2C and supplementary fig. 9).

In summary, brain impedance rises throughout the day during wakefulness, reaching a maximum in the evening before sleep when impedance cyclically decreases between states of NREM and REM sleep reaching an overall minimum in the early morning around wakening (Fig. 2C. supplementary fig. 8). Impedance is lowest in NREM, intermediate in REM, and highest during evening wakefulness (Fig. 1E & F). The brain state dependence of impedance and transitions between brain states generate impedance cycles with characteristic oscillations ranging between hours and days with a stable, dominant 24-hour cycle (Fig. 2). We propose these findings represent normal brain physiology given this analysis was focused on brain regions least involved in the subject’s epilepsy and only for days without seizure activity (Supplementary fig. 6). The variability of NREM and REM sleep duration and state transitions observed over the multiple month ambulatory recordings during the night underlies the ultradian cycle variability, and may be amplified here by the well-known sleep fragmentation in people with epilepsy^18^. We did not find a behavioral correlate for the infradian impedance cycles, but similar long-term cycles in physiologic and pathologic brain electrical activity are well known^19^.

The mechanisms underpinning the complex temporal dynamics and behavioral state dependent impedance cycles cannot be definitively determined from the data presented here. But we postulate that ECS volume changes drive the impedance cycles in humans observed here, consistent with the ECS volume expansion reported in rodent NREM sleep ^8,11^ and the established ECS dependence of electrical impedance^3,7^. Our data provide the first indirect evidence that behavioral state (wakefulness, NREM and REM sleep) differentially modulates human ECS, and is consistent with the ECS dynamics of the brain glymphatic system. The vascular and intracellular compartment conductance have limited contributions to composite impedance due to the high impedance of the blood brain barrier and neuronal and glial membranes. The electrode-tissue interface contribution to the impedance is not expected to depend on behavioral state, and should not differentially contribute to behavioral state related impedance changes. This is further supported by the persistence of circadian impedance oscillations in stimulation electrodes after periods of electrical stimulation that has been reported to disrupt the glia sheath around the electrode (Supplementary fig. 4). The interstitial fluid filled ECS at the known physiological volume fraction (∼0.2) and complex cellular geometry^7^ provides a highly conductive network that may be near the percolation threshold^20^ enabling significant impedance changes with relatively small ECS volume changes^20^. For these reasons, we propose ECS volume dynamics likely underlie the observed behavioral state dependence of impedance and the multiscale cycles. A less likely, alternative hypothesis is that a sleep state dependent change in astrocyte membrane impedance and gap-junction mediated astrocytic syncytium network provides a low impedance pathway^3^, and cannot be excluded with the current results. This seems less likely given the recent 2-photon imaging findings in rodents^8,11^.

This study has limitations in understanding the role of the ECS in physiological impedance oscillations: 1) The data are from people with focal epilepsy. Despite our selection of brain regions and periods without seizures there is electrophysiological evidence the brain regions analyzed are not entirely normal. In particular, as is generally the case in limbic epilepsy there are IES throughout the limbic network in all subjects. Nonetheless, here we exclude periods and electrodes generating the seizures. It is notable that the impedance fluctuations in the days with seizures, which are excluded in our analysis, show significant impedance variation compared to days with seizures (Supplementary fig. 6). This is a current area of investigation and suggests that epileptic brain impedance is changed at baseline. 2) The two-point impedance measurements introduce electrode-tissue interface effects and frequency dispersion (Supplementary fig. 5). The fact that high frequency electrical stimulation decreases baseline impedance without effecting the presence or characteristics of circadian impedance cycles (Supplementary fig. 4) supports the argument that impedance cycles are related to bulk tissue impedance changes and not changes at the electrode-tissue interface. 3) The claim that ECS dynamics underlie the impedance oscillations is not directly proven with the current data. In the future, we plan to use high temporal resolution impedance sampling combined with 2-photon imaging in animals to investigate impedance, ECS dynamics and the cycles of brain excitability observed in interictal epileptiform spike rates and seizures^19^.

## Data Availability

The data and code for analysis that support the findings of this study are available from the corresponding author upon reasonable request.

## Abbreviations

(AMG): Amygdala
(ANT): Anterior Nucleus of Thalamus
(EBS): Electrical brain stimulation
(ECS): Extracellular space
(HPC): Hippocampus
(INSS): Implantable Neural Sensing and Stimulation device
(*Z*): Brain impedance
(<Z>): Z-score of impedance
(LFP): Local field potentials
(REM): Rapid-eye-movement
(NREM): Non-rapid eye movement sleep

## Methods

### Human subjects

Five people with drug resistant bilateral mesial temporal lobe epilepsy were implanted with an investigational neural sensing and stimulation device under FDA IDE: G180224 and Mayo Clinic IRB: 18-005483 *Human Safety and Feasibility Study of Neurophysiologically Based Brain State Tracking and Modulation in Focal Epilepsy*. The patients provided written consent in accordance with IRB and FDA requirements. Study registration https://clinicaltrials.gov/ct2/show/NCT03946618.

#### Protocol & Experimental Design

Subject inclusion required demonstration of drug resistant^21^ mesial temporal lobe epilepsy (mTLE). For inclusion the patients had bilateral independent left and right temporal lobe onset seizures, or seizures from the dominant temporal lobe. Patients were required to have three or more disabling seizures per month as demonstrated on a mobile epilepsy patient assistant application (EPA)^22^ diary. Patients were required to have disabling focal aware seizures (FAS), focal impaired awareness seizures (FIAS) or focal to bilateral tonic-clonic seizures (FBTC)^23^. Nine patients were consented and five successfully completed the seizure diary baseline and were implanted with the investigational Medtronic Summit RC + S™ device (Supplementary fig. 1). Patients remained on stable medication regimes over the course of the study.

#### RC+S™ Summit System (Supplementary figure 1)

The local field potential (LFP) and impedance data were acquired using the RC+S™ Summit. Each patient had four implanted leads targeting left and right anterior nucleus of thalamus (ANT), amygdala (AMG) and hippocampus (HPC)^13,15,24^.

### Lead and Electrode Contact Localization (Supplementary fig. 2)

Bilateral ANT (Medtronic 3387 leads) and HPC & AMG (Medtronic 3391 leads) were stereotactically implanted in 4 subjects. The 16 electrode contacts were localized with post-operative CT co-registered to the pre-op MRI for anatomic localization using previously described pipelines^25,26^. The CT and electrode contact positions are co-registered to a T_1_ weighted anatomical MRI scan using co-registration in SPM12 (https://fil.ion.ucl.ac.uk/spm/)^27^. Freesurfer (http://surfer.nmr.mgh.harvard.edu/) was used to segment the T_1_ MRI and the electrodes labeled according to the Destrieux atlas ^28,29^. The final electrode contact localization for impedance analysis was performed with Lead DBS^26^ (Supplementary Fig. 2).

*The RC+S*^*TM*^ *Summit* device *(Supplementary fig. 1)* enables programmable16 channel electrical stimulation, monopolar and bipolar 2-point impedance measurements, and continuous 4 channel (selected bipolar pairs) wireless streaming of brain local field potentials (LFP) to a handheld tablet computer and cloud environment^15,22,30^. The integration and synchronization of the RC+S™ Summit with off-the-body computing devices (tablet computer, smart phone, watch) uses a custom software application (Epilepsy Patient Assistant: EPA)^15,22,24^. The RC+S™ requires ∼1.5 hours of charging for continuous 24-hr electrical brain stimulation (EBS), and LFP and impedance timeseries wireless streaming to the handheld tablet. The telemetry antenna requires charging every 72 hours. The tablet computer running the EPA application is maintained with wall power and has ∼8 hours rechargeable battery.

### Sensing Local Field Potentials (LFP)

The LFP signals were continuously recorded for 2,308 days (M1 - 828 days; M2 – 549 days; M3 - 346; M4 – 549; M5 – 36 days) while trialing several EBS paradigms applied to the ANT^13,24^. The LFP signals were recorded with sampling frequency of 250 Hz or 500 Hz. The sampling frequency was determined as a compromise between data quality, wireless data transmission dropout, and device battery limitations.

### Automated LFP Analysis of Epileptiform Activity and Behavioral State

We deployed a previously developed suite of automated tools for sleep classification^13^, interictal epileptiform spike (IES) and seizure detection^24^ synchronized with patient reports of seizure, mood and sleep using the EPA^22,31^. The system (Fig. 1.) is integrated with a cloud computing environment providing visualization of continuous LFP data, brain state classification, IES rates, seizures, and the information entered by patients. The automated electrographic seizure detection algorithm running on the tablet and in the cloud identifies electrographic seizures using high sensitivity thresholds. The candidate seizure events are subsequently visually reviewed and confirmed or rejected by a human expert.

### Behavioral state classification

We used a previously validated algorithm developed from simultaneous polysomnography (PSG) and LFP recordings^13^. Expert visual review was used to create gold standard labeled training, validation and testing data as defined by AASM^32^: Awake, Rapid-eye-movement (REM) sleep, and three Non-REM (N1-N3) sleep stages. The automated classifier using LFP signals recorded from AMG, ANT, and HPC produced accurate classification (average F1-score 0.89 on testing data) and was subsequently deployed in the five patients for behavioral state classification (Awake, REM, Non-REM (N2 & N3)). *Naps:* Four subjects took sporadic naps making impedance analysis of slow wave sleep during naps possible. The periods of nap N2 & N3 sleep (> 30 min. duration) were compared to wakefulness in ANT, AMG and HPC (Fig.1).

#### Electrical impedance measurements (Supplementary fig. 3)

We sampled electrical impedance from 16 electrodes (AMG, HPC and THL) every 5 - 15 minutes. The impedance was measured for each electrode contact using a 2-point measurement between the electrode contacts and the RC+S™ device (monopolar impedance) and between bipolar electrode contact pairs using a single charge balanced square-wave current pulse (0.4 mA, 80 μsec pulse width). For the 2-point impedance measurement configuration the same electrode contacts are used for delivering the current stimulation and sensing the voltage response. The measured impedance, 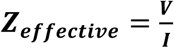, is determined from the measured voltage (***V***) and injected current (***I***). The contributions to the effective impedance include the two electrode contacts, electrode-tissue interface, and the bulk brain tissue. The brain tissue impedance includes the extracellular space, intracellular space, and vasculature space contributions.

### Brain tissue reaction to implanted electrodes and electrical stimulation (Supplementary fig. 4)

To avoid the effect of tissue injury on brain impedance measurements from the implant and immunologic foreign body response we restricted the data analysis to greater than 14 days post-implantation. The response to implanting an electrode foreign body includes the acute local tissue injury followed by immunologic response of macrophages, giant cells, glia and accumulation of collagen and fibroblasts around the electrode. The resulting sheath of encapsulation tissue around the electrode has a dynamic, frequency dependent effect on impedance^33^. The impedance reaches a stable maximum in 7 – 14 days in animals^34^ and humans^12,35^. It is important to account for the early changes related to encapsulation tissue that will alter the impedance measurements and electric fields generated by implanted electrodes. Interestingly, electrical stimulation can be used to decrease impedance^36,37^ by disrupting the tissue encapsulation. For these reasons we have not included electrodes used for EBS in this study.

#### Saline/microbead Impedance and RC+S™ measurements (Supplementary fig. 5)

We performed benchtop experiments in saline and saline/microbead composites to directly compare 2-point and 4-point electrode impedance measurements using sinusoidal currents with single pulse RC+S™ impedance measurements. The saline/microbead composite is used to model brain extracellular space and conductive interstitial fluid (Supplementary Fig. 5B).

### 2-point and 4-point electrode contact Impedance Measurements (Supplementary fig. 5A)

The investigations used a Medtronic 3387 lead (4 contacts each with 4.7mm^2^ surface area) submerged in a plastic reservoir (1 cm *x* 5 cm x 1 cm) of saline (0.9% Sodium Chloride solution at 22°C; 68.9Ω-cm)^38^ or saline/microbead composites (volume fractions 36% saline and 64% microbeads (80 -120 μm))^39,40^. To simulate monopolar electrical impedance measurements a single electrode contact on the 3387 lead was the cathode and a large surface area contact was the anode (modeling the large RC+S™ surface area ∼ 5.4×10^3^ mm^2^). Given the large surface area of the RC+S™ device the impedance is small compared to the 3387-electrode contact, electrode-tissue interface and local environment. For 2-point bipolar measurements the inner 2 contacts of the 3387 were used for stimulation and recording. For 4-point measurement the outer 2 contacts of the 3387 were used for injecting stimulation current and the inner 2 contacts used for recording the voltage. A National Instruments USB-6251 signal generator and a NPI electronics ISO-isolation unit (www.npielectronic.com) were used to deliver 0.5 μA sinusoidal currents over a wide frequency range (1 – 1500 Hz). The voltage drop across the measurement electrodes was recorded using a Neuralynx-Cube acquisition system (www.neuralynx.com). The 2- and 4-point impedance measurements were directly compared to RC+S™ measured single pulse (0.4mA, 80us pulse width) impedance measurements (Supplementary fig. 5C).

The frequency dispersion observed in the 2-point measurement is generated by the capacitive double layer^41,42,43^ that forms at the electrode-tissue interface. The 4-point impedance measurement uses different electrodes for delivering stimulation and measuring voltage, and shows no frequency dispersion by eliminating the stimulation induced polarization at the electrode-media interface^44^ (Supplementary figs. 5A&C). We directly compared 2-point & 4-point impedance measurements using sinusoidal current and RC+S™ impedance measurements in saline and saline/microbead composite. The electrical impedance measured using the RC+S™ charge balanced square-wave pulse corresponds approximately to the 2-point measurement at ∼2 kHz sinusoidal current in both saline and the saline/microbead composite, and is ∼10x higher than the 4-point sinusoidal measurements (Supplementary fig. 5C). We also evaluated the effect of temperature on RC+S™ impedance measurements over the physiologic range of wakefulness and sleep temperatures. With the RC+S™ and a 1.2 kΩ resistor sealed in a temperature-controlled environment there was no detectable impedance change between 32°C and 40°C.

#### Electrical Brain Impedance Analysis in Human Subjects

We acquired brain impedance (sampling rate 5 to 15 minutes) over the course of 1382 days with the impedance 24-hour data rate greater than 70% (M1 – 579; M2 – 55; M3 – 401; M4 – 338; M5 – 9 days). The impedance time series were initially resampled with a 30-minute long moving average window with step of 10 minutes and bandpass-filtered between 1 hour to 100 days with respective low- and high-pass finite impulse response (FIR) filters of 1,001^st^ order (Fig. 1C & supplementary figs. 3, 5-7).

We utilized 36 electrodes from right hemisphere AMG, ANT and HPC in the 5 subjects to analyze electrical brain impedance since the right mesial temporal structures showed significantly lower IES rates and seizures p<0.05. Only data that removed 24 hours from seizure activity is used in the impedance analysis. (Supplementary figures 2 & 6).

Electrophysiology confirmed seizures (paired t-test)

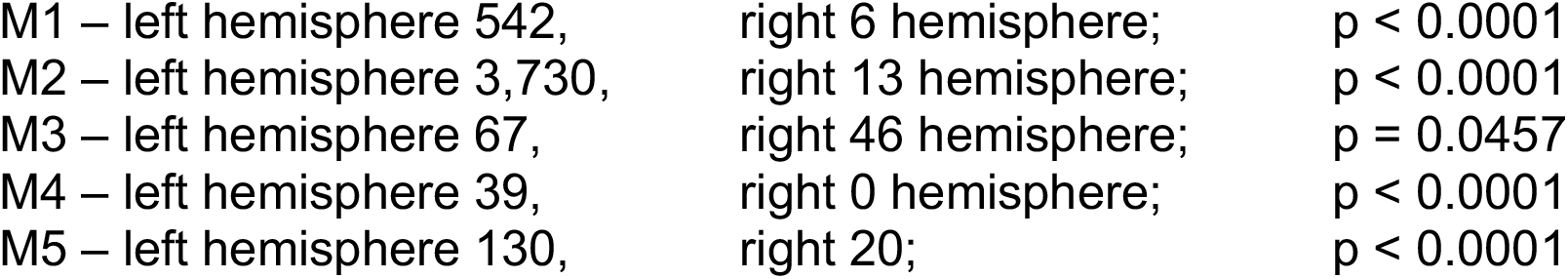

|  |  |  |
| --- | --- | --- |
| M1 – left hemisphere 542, | right 6 hemisphere; | $p < 0.0001$ |
| M2 – left hemisphere 3,730, | right 13 hemisphere; | $p < 0.0001$ |
| M3 – left hemisphere 67, | right 46 hemisphere; | $p = 0.0457$ |
| M4 – left hemisphere 39, | right 0 hemisphere; | $p < 0.0001$ |
| M5 – left hemisphere 130, | right 20; | $p < 0.0001$ |

Interictal Epileptiform Spikes rates (IES/day) (paired t-test)

|  |  |  |
| --- | --- | --- |
| M1 – left $1,941.4 \pm 889.8$, | right $132.8 \pm 212.9$; | $p < 0.0001$ |
| M2 – left $1,924.4 \pm 845.5$, | right $588.2 \pm 455.5$; | $p < 0.0001$ |
| M3 – left $1,122.7 \pm 665.8$, | right $521.5 \pm 282.4$; | $p < 0.0001$ |
| M4 – left $140.6 \pm 189.8$, | right $22.3 \pm 55.9$; | $p < 0.0001$ |
| M5 – left $831.9 \pm 805.3$, | right $339.4 \pm 493.6$; | $p < 0.0001$ |

### Impedance and Seizures (Supplementary fig. 6)

Brain tissue impedance is reported to be transiently effected by seizure activity in animal models^17^. In a cat chemotoxin (tungstic acid gel) model of focal epilepsy and HPC seizures a regional impedance change was reported in ventral HPC (10 – 12%), modest change in dorsal HPC and AMG, and no change in brain areas not involved in seizure propagation. Impedance changes began after seizure onset and relatively quickly (20 – 90 seconds) returned to baseline after seizure offset^17^. However, early invasive investigations in human temporal lobe epilepsy did not see any impedance change with spontaneous seizures^45^. To minimize the potential effect of pathological epileptogenic tissue and seizures here we focused the impedance analysis on electrodes with low IES rates and data segments without any seizures within 24 hours. Interestingly, there is a relatively subtle difference in impedance in the 24-hr data epochs with seizures versus epochs without seizures and this is currently an area of investigation (Supplementary fig. 6).

### Impedance and Behavioral State (Wakefulness, NREM & REM Sleep (Supplementary figure 7)

In total 299 data segments from the 36 right hemisphere electrodes without seizure activity for at least 24-hours were annotated across the 5 patients and 3 categories Awake, NREM & REM (M1: 30/21/22, M2: 20/19/18, M3: 15/14/13, M4: 22/27/29, M5: 12/19/18). We compared three categories on a group and subject level using statistical two-tailed Mann-Whitney test (p < 0.05, Bonferroni correction for multiple observations). The data shows strong relationships between the electrical brain impedance in Awake, NREM & REM sleep states in all 5 subjects (Fig. 1 & supplementary fig. 7). The electrical brain impedance was significantly decreased in NREM compared to wakefulness and REM in multiple limbic brain targets (HPC, AMG, ANT). Note M5 only had data from HPC & ANT electrodes. Interestingly, ANT REM impedance is increased compared to NREM but remains significantly lower than wakefulness impedance at a group level, and individually three patients (M1-3) with consistent trend for M4 and M5.

### Impedance during naps

Four subjects took sporadic naps making impedance analysis of sleep during naps in addition to the overnight sleep/wake cycles possible. The continuous behavioral state classifications were reviewed for periods of sleep (> 30 min. duration). The Impedance was analyzed during and around these periods of non-REM sleep and compared to wakefulness in HPC, AMG and ANT channels (Fig. 1D). We have utilized the same preprocessing and pipeline for 18 daytime naps identified during the monitoring period distributed across 4 patients (M1: 7, M2: 1, M3: 7, M4: 3). An average impedance value during a daytime nap with duration of at least 30 minutes was compared to average impedance of 1 hour pre-nap period using two-tailed paired Wilcoxon test. The impedance was reduced in HPC and AMG in slow wave NREM sleep compared to 1-hour pre-nap wakefulness period (p < 0.05). The result supports that the impedance cycles are related to behavioral state changes, and not directly related to a circadian cycle mechanism.

#### Multiscale Cycles of Brain Impedance

We investigated ultradian (< 24-hr), circadian (∼24-hr) and infradian (> 24-hr) cycles in brain impedance (Fig. 2 and supplementary figs. 8 & 9). Analysis of the magnitude of the min-max impedance difference within 24-hour cycles for AMG, HPC and ANT was 46.06 ± 16.72 Ω, 46.43 ± 15.72 Ω and 48.94 ± 16.43 Ω, respectively. The the 24 hour cycle histograms are nonuniformly distributed (circular OMNIBUS^46^) reflecting the evening maximum impedance occurring at 7:04 PM ± 3:55 hr. and morning minimum impedance at 6:15AM ± 1:56 hr. (Kuiper two-sample test ^47^).

### Brain impedance (*Z*) *spectral analyses*

Continuous wavelet transforms (CWT) implemented in MATLAB with Morlet wavelets^48^ (L2 normalization) and minimum/maximum scales determined by the energy spread of the wavelet in frequency and time were used to investigate the presence of multiscale impedance rhythms (Fig. 2A). For significance testing of putative multiscale impedance cycles a 5-day moving window (moving step of 5 days) was used to test significant ultradian and circadian cycles and a 100-day moving window (moving step of 5 days) significant infradian cycles (Supplementary Figure *9)*. Testing for ultradian and circadian band cycles for each 5-day signal segment was investigated using Thomson F-test multi-taper scheme^48,49^ to test the periodicity at the frequencies of interest (FOI, Supplementary fig. 9).

Periodicity test in infradian band used the same procedure, except that the window length was set at 100 days and FOI was from 2.5 days/cycle to 30 days/cycle with an incremental step of 15 minutes. (Supplementary fig. 9)

**Supplementary figure 1.**
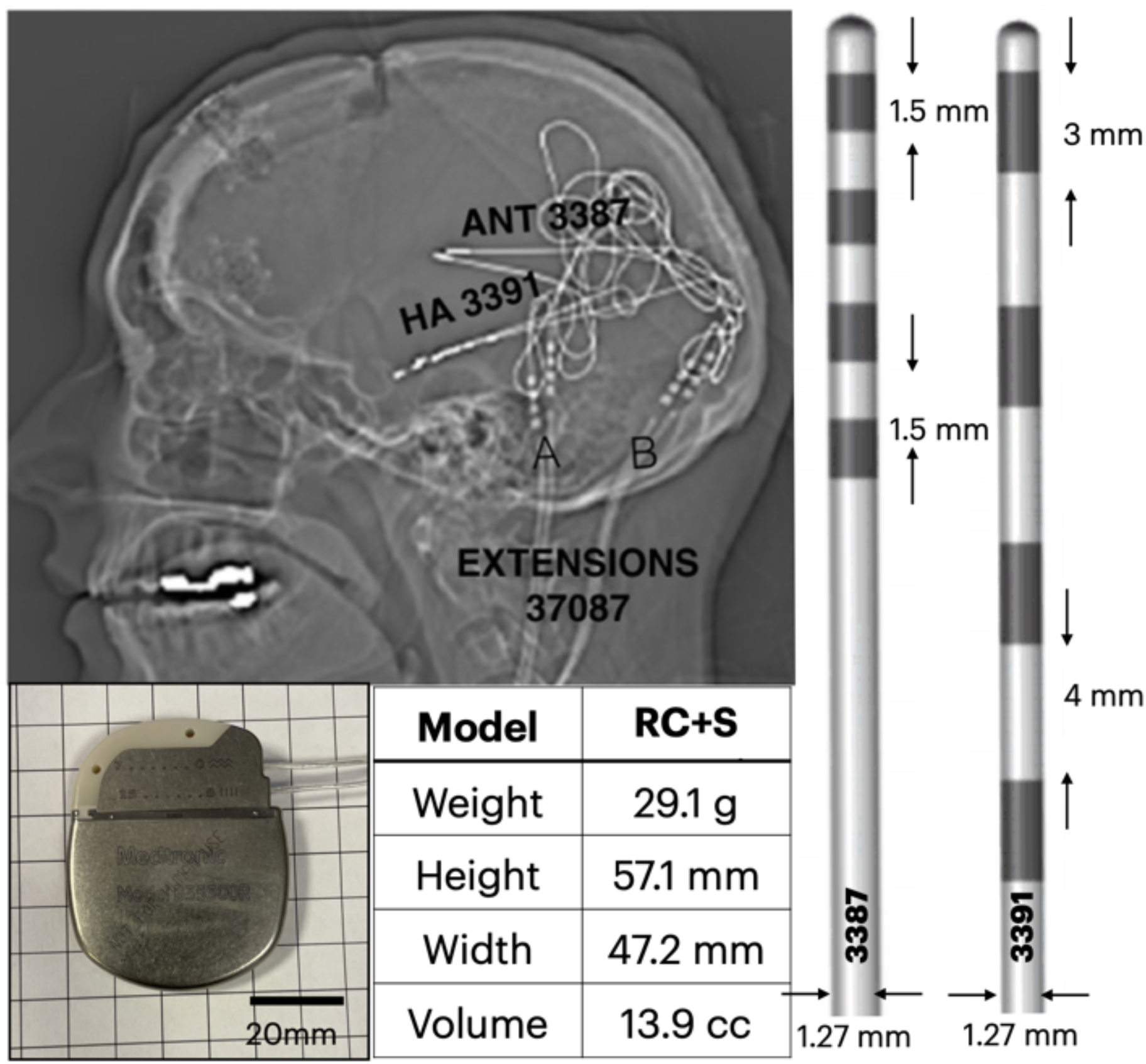
Limbic Network Implant for Mesial Temporal Lobe Epilepsy (mTLE). The Medtronic RC+S™ enabled continuous tracking of limbic network electrophysiology and behavior. **A)** Lateral x-ray (M1) showing 3387 electrodes in the bilateral ANT and 3391 electrodes in bilateral AMG and HPC. **B)** The 3387 four contact (contact surface area = 5.985 mm^2^) lead span 10.5 mm. The contacts are 1.5 mm long each and separated by 1.5 mm. The 3391 four contact (surface area = 11.97 mm^2^) lead span 24.5 mm. The contacts are 3.0 mm long and separated by 4.0 mm. **C)** RC+S™ Summit implantable device dimensions. The rechargeable device enables 16 stimulation channels and programmable 4 sensing channels from any bipolar contact pair.

**Supplementary figure 2.**
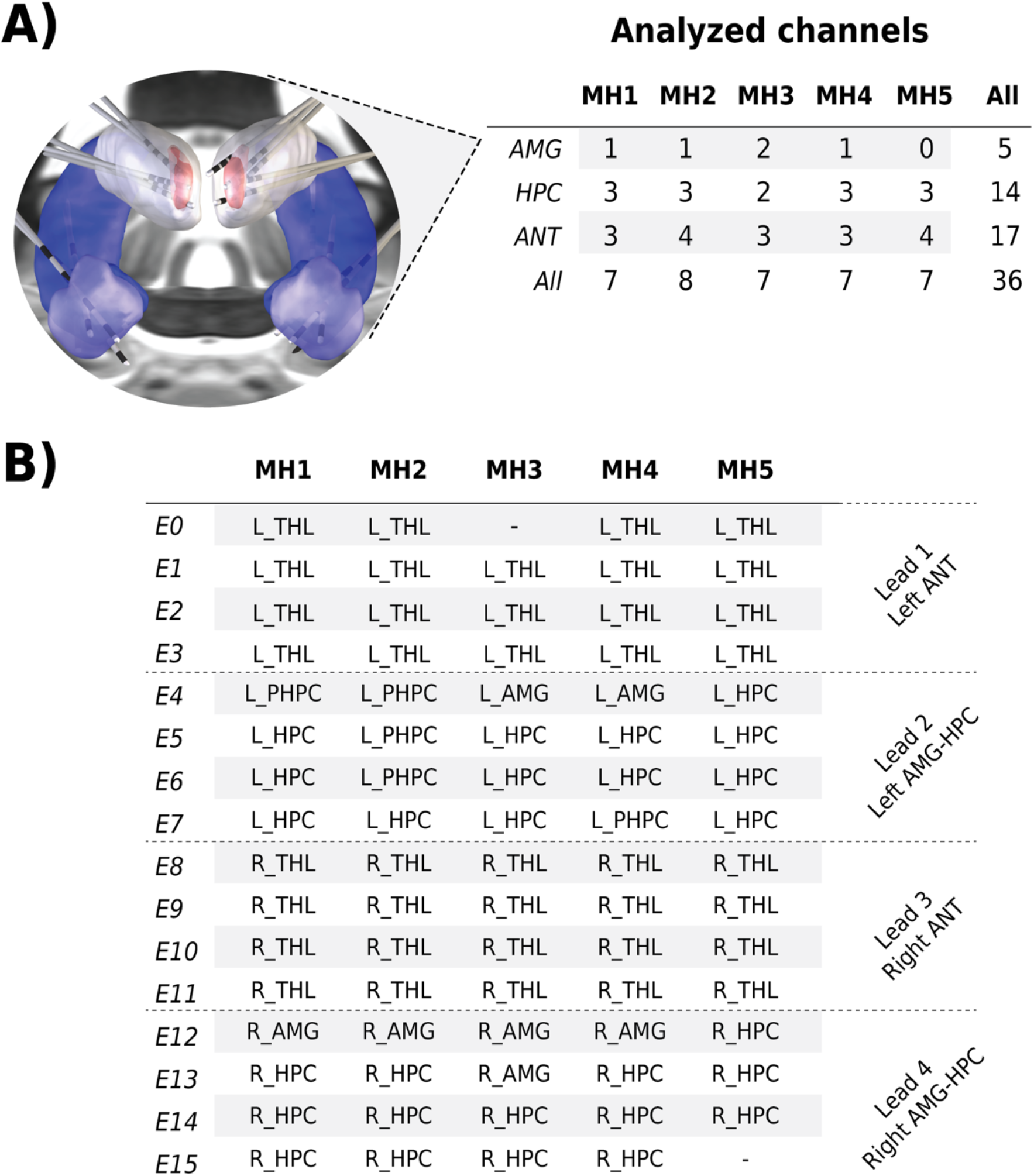
Electrode Contact Localization. Multiple limbic network nodes were targeted. **A)** Lead DBS localization of bilateral Amygdala and Hippocampus (AMG & HPC: Yellow), Anterior Nucleus Thalamus (ANT: Red) leads and electrode contacts. There were 5 AMG, 14 HPC and 17 ANT right hemisphere contacts used for impedance analysis. **B)** Electrode Contact Localization to Pre-Op MRI. Left (L) and Right (R) thalamus (THL), amygdala (AMG), hippocampus (HPC) and parahippocampal gyrus (PHPC).

**Supplementary figure 3.**
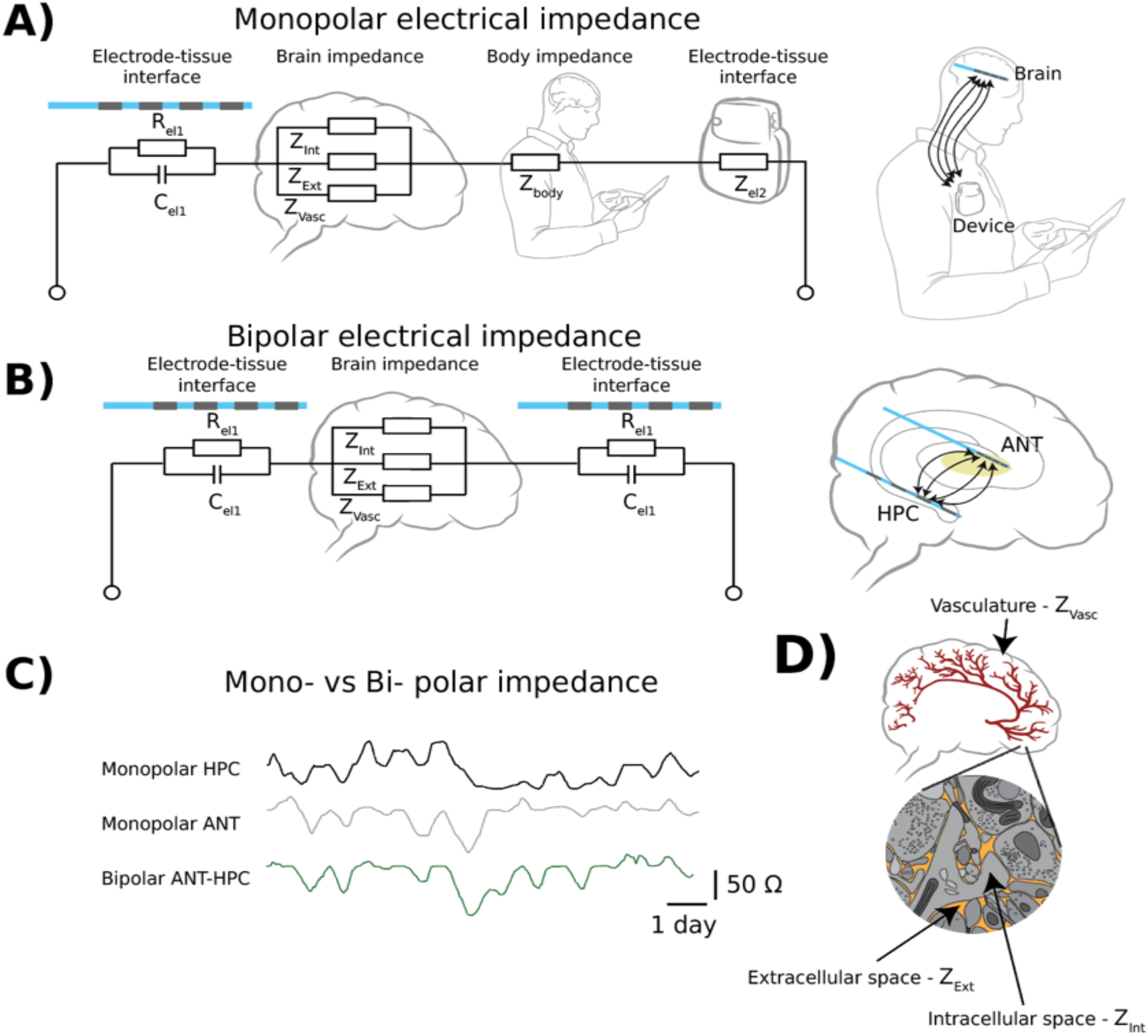
Human Brain Impedance. Electrical 2-point monopolar and bipolar impedance measurement were periodically sampled in 5 subjects. **A)** The 2-point monopolar measurement injects a local current into the brain using one of the 3387 (surface area=5.98^2^) or 3391 (surface area = 11.97 mm^2^) contacts as the cathode (E_1_). The current return (anode (E_2_)) is the low impedance RC+S™ device (surface area 5.4×10^3^ mm^2^) in the chest sub-clavicular pocket. **B)** The 2-point bipolar configuration uses two different 3387/3391 electrode contacts as cathode and anode. Right) Circuit schematics for the 2-point monopolar and bipolar impedance measurements. **C)** The monopolar impedance is ∼ 50% less than the bipolar impedance given the RC+S™ Summit surface area is ∼10^3^ greater than the 3387/3391 electrodes. **D)** The schematic of brain microenvironment composed of cellular, extracellular, and vascular compartments.

**Supplementary figure 4.**
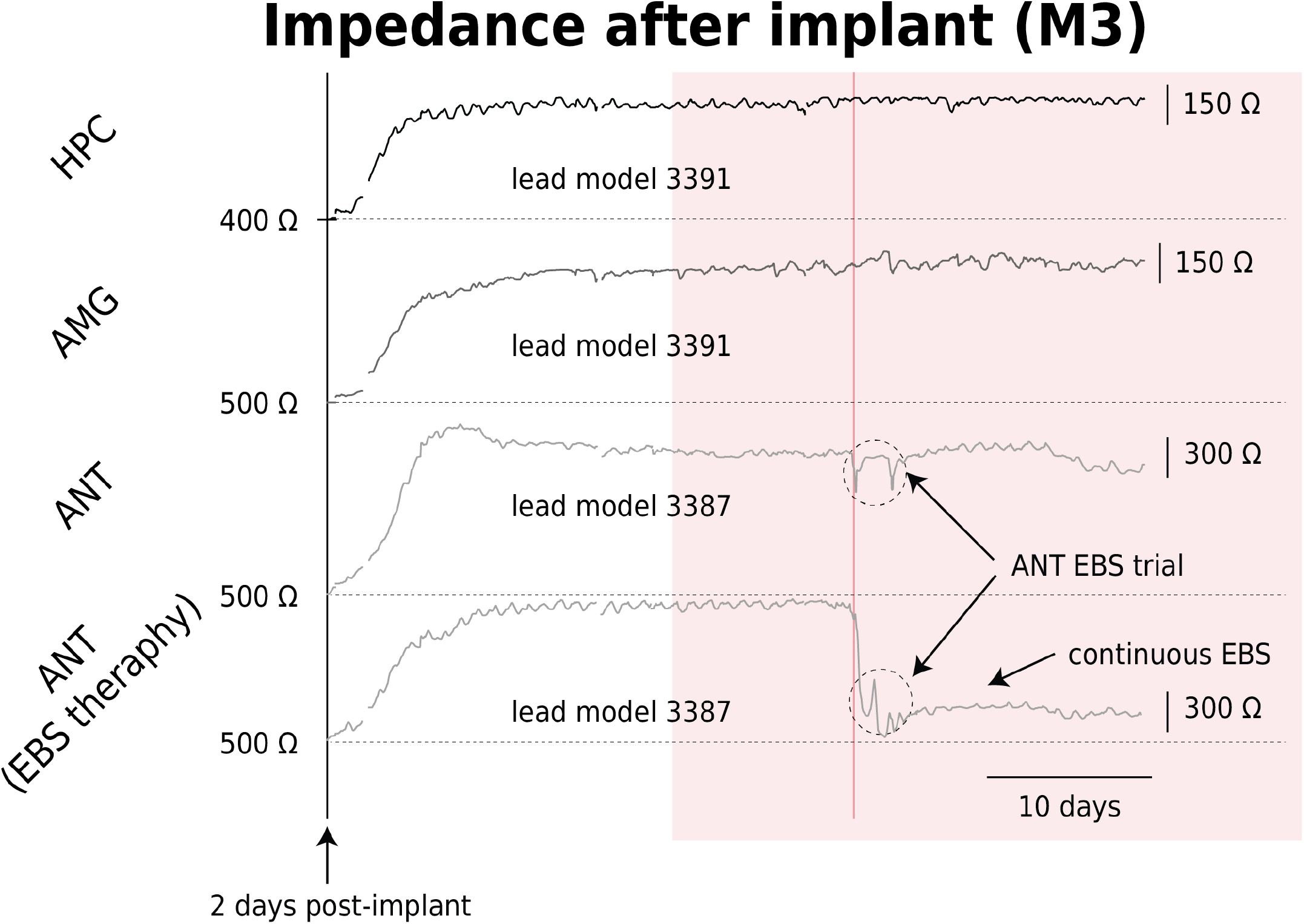
Long-term Brain Impedance and Electrical Stimulation. Electrode-tissue impedance and foreign body gliosis around the electrode stabilizes over the 15 days after implant, but is rapidly and reversibly changed with electrical stimulation. Top) HPC and AMG impedance over initial 2 months after implant. After implant the impedance increases over the next ∼14 days to a stable mean value. Bottom) The ANT impedance exhibits similar behavior to HPC and AMG. The application of electrical brain stimulation (EBS) to ANT electrodes produces an immediate drop in impedance to a new stable minimum. Note the circadian cycles of impedance can be appreciated in AMG, ANT and HPC.

**Supplementary figure 5.**
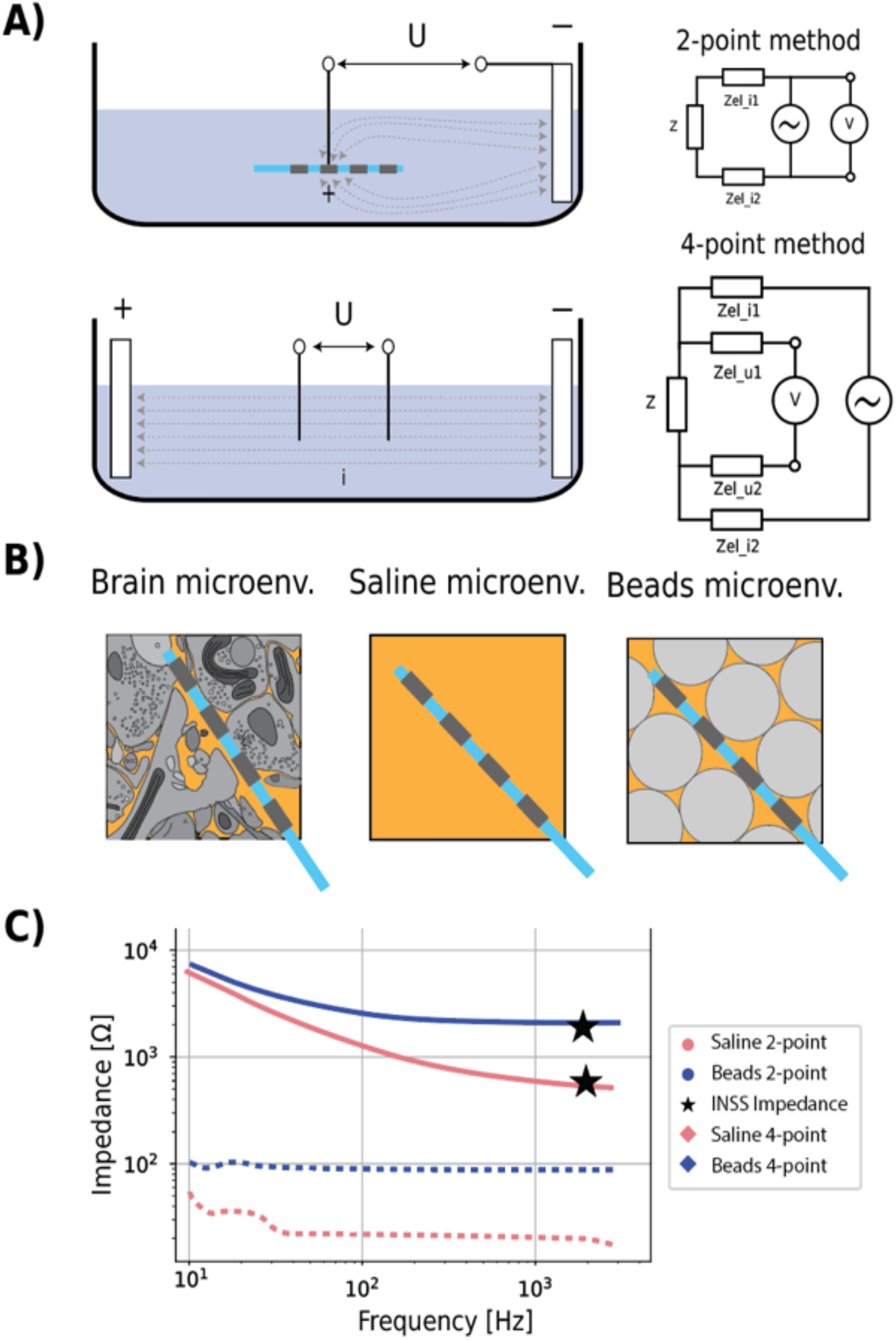
Impedance Measurements in Saline and Saline/Microbead Composites. Benchtop testing of 2- & 4-point impedance measurements was performed and compared to RC+S™ measurements. **A)** For 2-point monopolar measurements current is injected into the sample medium using one of the 3387 contacts as the cathode (E_1_) and the anode (E_2_) is a low impedance, large surface area contact. The 2-point bipolar measurement uses the same electrodes (E_1 &_ E_2_) for both electrical stimulation and voltage sensing. The 4-point impedance measurement uses different electrodes for stimulation (E_1 &_ E_2_) and sensing (E_3 &_ E_4_). **Right)** Circuit schematics for 2-point and 4-point impedance measurements. **B)** Schematic of different media environments. 1) Brain microenvironment composed of cellular, extracellular, and vascular compartments. 2) Saline and, 3) Composite of microbeads and saline. **C)** Impedance (10 – 1500 Hz) measured in saline and saline/microbead composites. The 2-point measurements are dominated at low frequency (< 10 Hz) by the frequency dependent capacitive double-layer. For the 4-point impedance measurement utilizing different electrodes for the current injection and sensing the impedance is not frequency dependent. The impedance measured with the RC+S™ device (⋆) using the pulse (0.4 mA, 80 μs pulse width) compared to sinusoidal currents show that the 2-point RC+S impulse response impedance is similar to a 2000 Hz sinusoidal current.

**Supplementary figure 6A.**
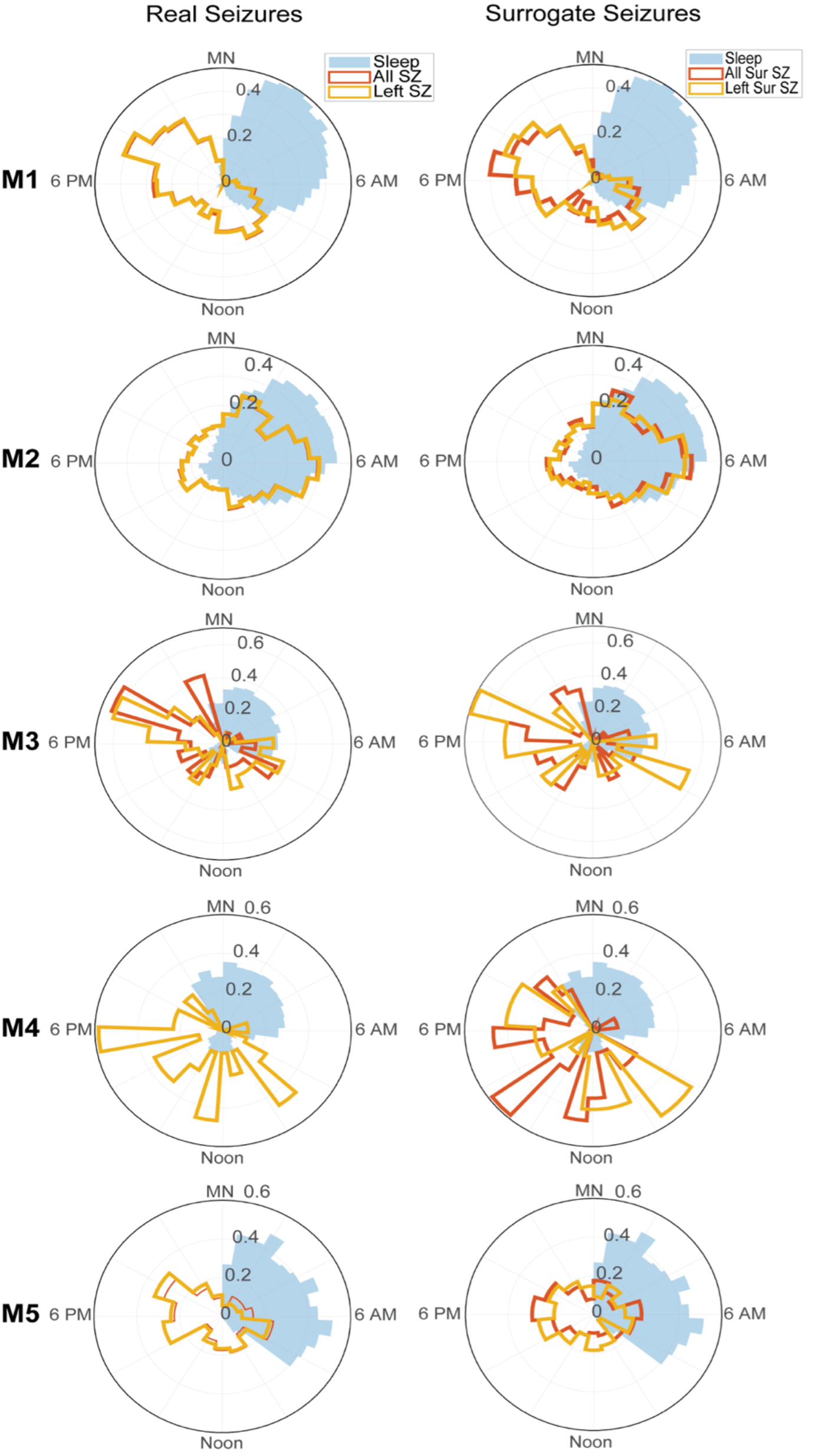
Seizure Probability over 24-hour period. Probability of seizure and sleep as a function of time of day (TOD). For each subject, the left panel shows the sleep probability (blue filled histogram bars) superimposed by the probability of *real* seizure onset times as a function of TOD. The red histograms indicate the probability of all types of seizures (left side, right side, both sides, and self-reported seizures), while the yellow histograms are left side seizures only (M1: number of all types of seizures / number of left seizures = 544 / 537, M2: 3758 / 3728, M3: 112 / 66, M4: 39 / 39, M5: 149 / 129). The right panel shows the sleep probability superimposed with the probability of *surrogate* seizure onset times. The surrogate times were generated for 24-hr. periods without any seizure activity to create equivalent distributions of times for comparing impedance around seizures with impedance without seizures. The surrogate distribution accounts for the natural circadian impedance cycles. The estimated probability distribution of real seizures is created using the “rejection method” based on the geometric interpretation of probability distribution ^50^. The red histograms indicate the probability of surrogate seizures of all types and the yellow histogram the surrogate left seizures. The number of generated surrogate seizures were the same as the corresponding types of real seizures.

**Supplementary figure 6B.**
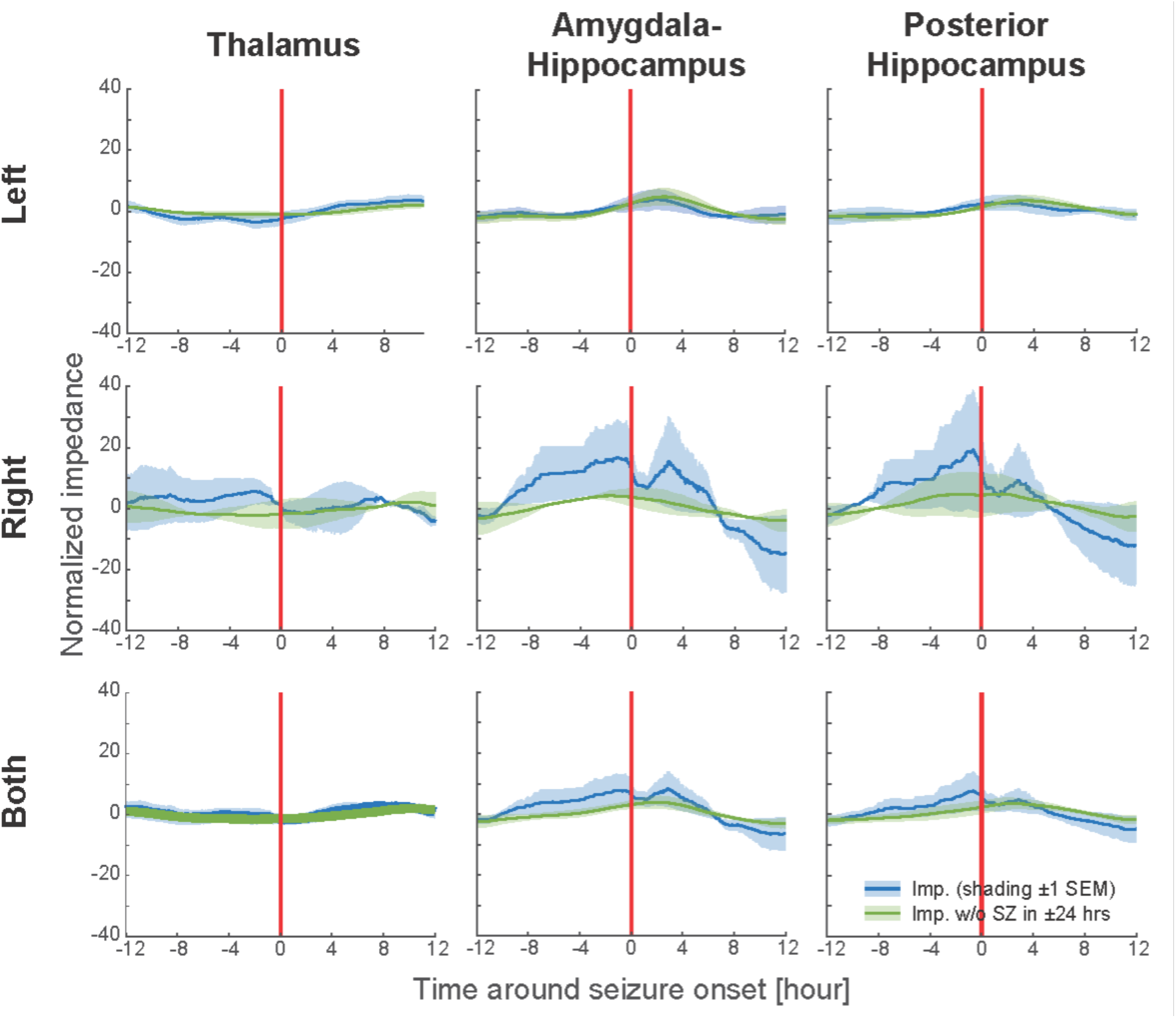
Seizure Related Impedance Change and Circadian4r. The impedance in three anatomical locations in limbic network (AMG, ANT, and HPC) was investigated in the 24 hours surrounding spontaneous seizures. Raw impedance times series were segmented ±12 hrs. around the seizure onset times. Left/right temporal lobe seizures were chosen for the impedance measured from left/right hemisphere limbic network structures. For each segment, the mean value of the impedance was subtracted to obtain the normalized impedance. All normalized impedance time-series were aggregated and smoothed with a moving window of 2.4 hrs. For each subject, the smoothed, normalized impedances were aggregated and the mean value per hemisphere location (Left, Right or Both sides) per anatomical location (Thalamus, Amygdale-Hippocampus and Posterior Hippocampus) was calculated. In each panel, red vertical line indicates the onset of seizures (for blue traces) or surrogates without seizures within 24 hours (green traces) that have same temporal distribution. The blue curve is the mean value of seizure-related normalized impedance, involving all available seizures and the green curve is the mean value of normalized impedance from 24-hour segments without any seizures within ±24 hrs. The timing of impedance data segments without seizures (green) are matched with seizure timing for time of day to account for any baseline circadian cycle changes. The shadings around the means indicate ±1 SEM, the uncertainty across the subjects (n = 4). **Abbreviations**: Impedance (IMP); seizure (SZ); standard error around mean (SEM); hours (hrs),

**Supplementary figure 7.**
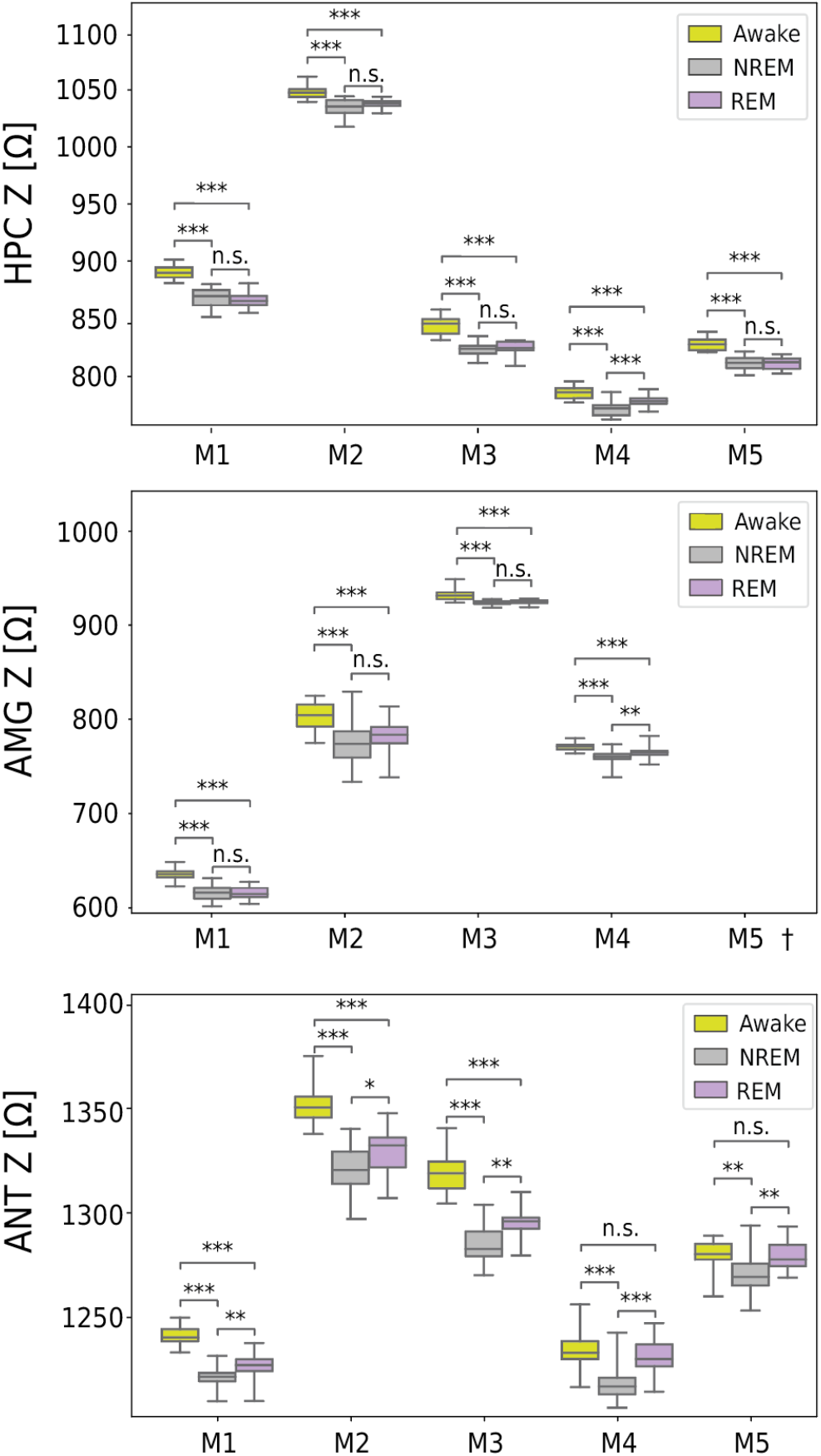
Brain Impedance Behavioral State Dependence. Comparison of behavioral state-dependent impedance in NREM, REM and Wakefulness. The AMG, ANT and HPC impedance is lowest in NREM, intermediate in REM and highest in wakefulness (Figure 1E). On the group level all behavioral state impedances are different in HPC, AMG and ANT. On the single subject level for HPC and AMG the impedance in NREM is lower than Awake. NREM impedance is significantly lower than REM for M4. For all subjects the ANT impedance is significantly lower in NREM v.s REM and NREM v.s Awake. But only 3 subjects (M1, M2 and M3) show significant difference between Awake and REM. The impedance difference between Awake and REM is smaller than what is observed between NREM and Awake or NREM and REM. This suggests that REM and Awake, activated states have more similar ECS volumes. (*p < 0.05, **p < 0.01, ***p < 0.001 Mann-Whitney test with Bonferroni correction for multiple observations). † AMG data were not available for M5.

**Supplementary figure 8.**
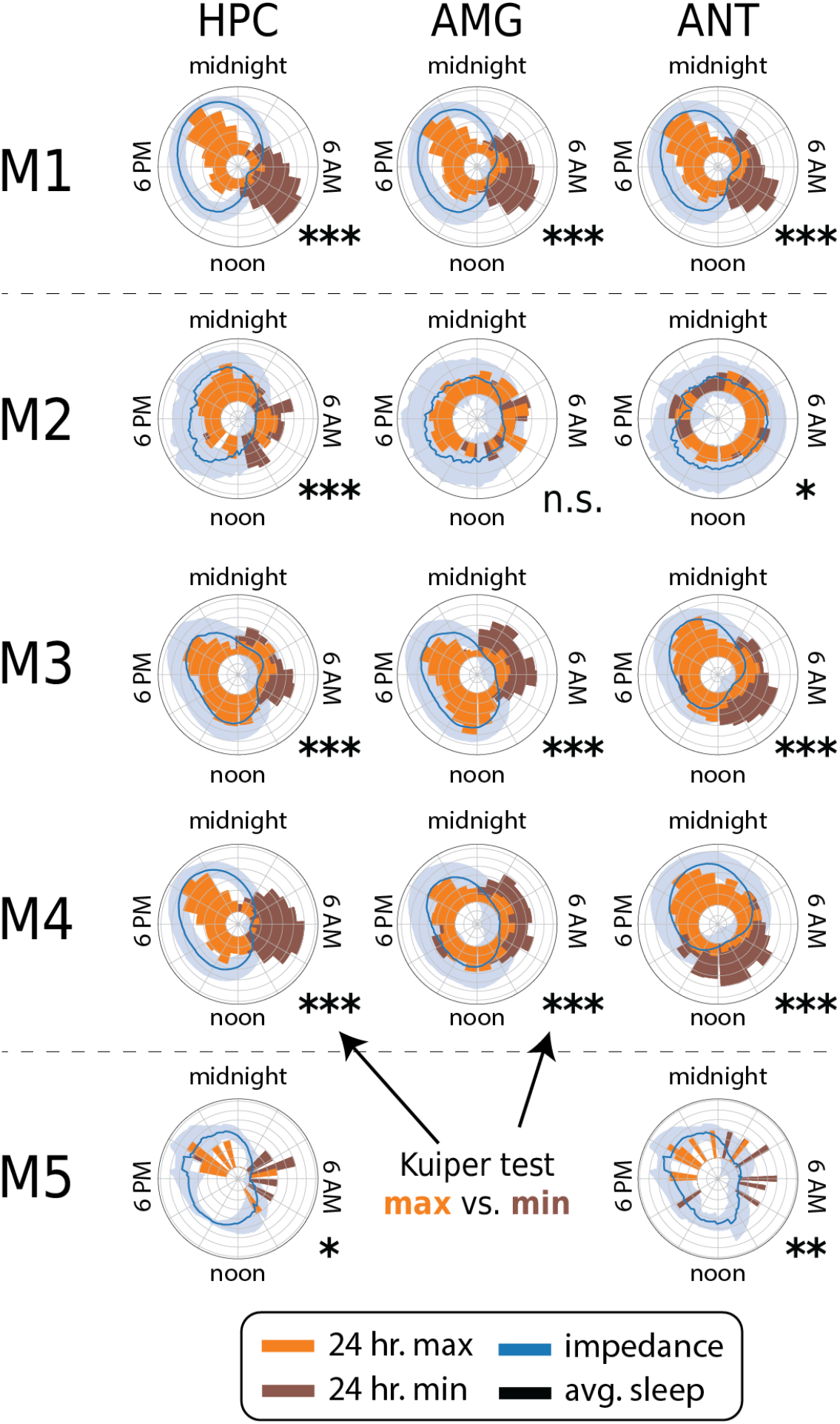
Circadian Impedance Cycles. Impedance signals were bandpass filtered between 6 hours and 30 hours using a 1001^th^ order of finite inpulse response, zero phase shift filters. Polar plots subjects M1-5 depicting a histogram distribution of the min and max values of the 24 hour impedance cycle. The minimum and maximum values were tested using circular Kuiper test (*p<0.05 ;**p < 0.01;***p<0.001).

**Supplementary figure 9.**
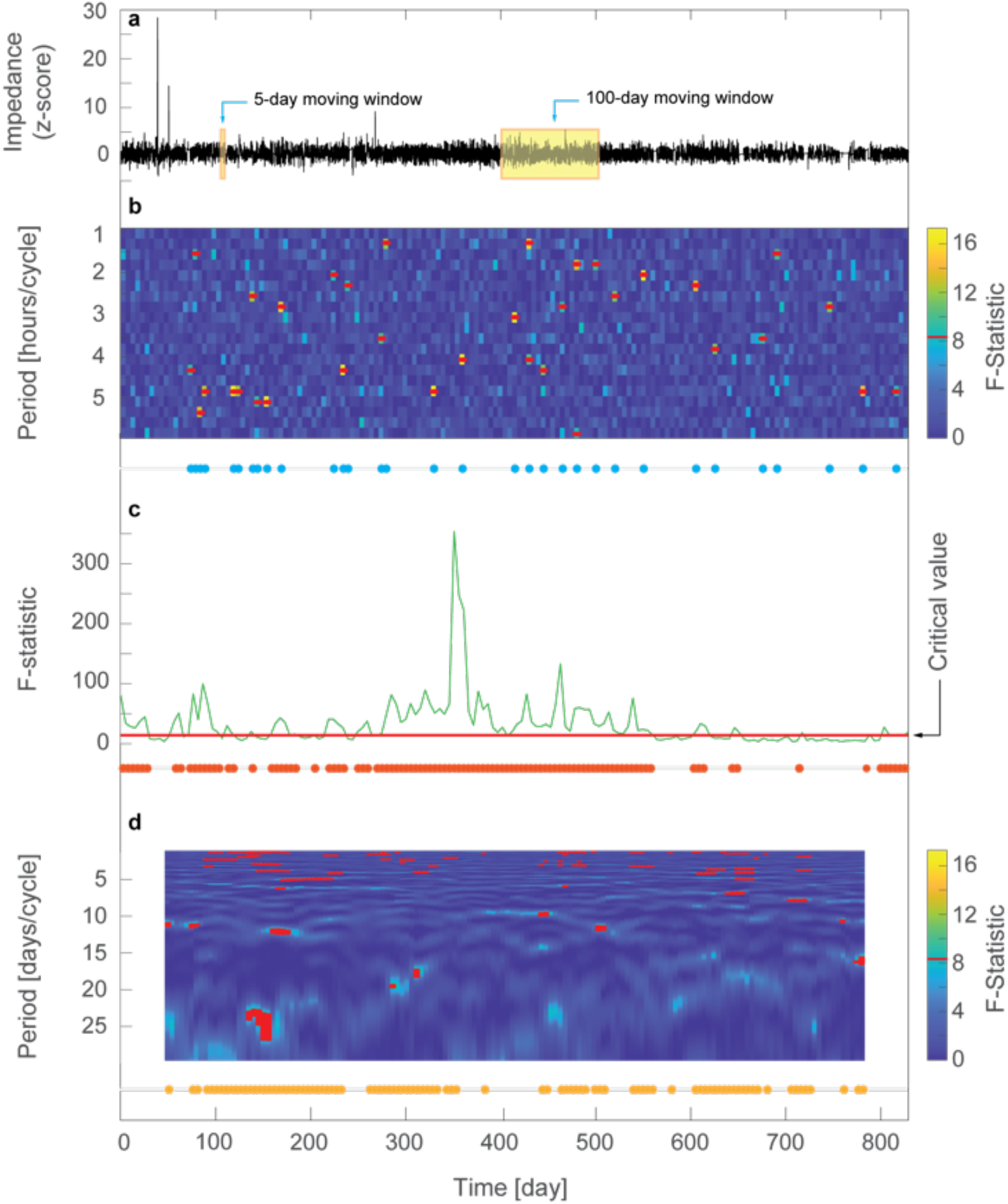
Periodicity tests for Impedance Oscillations. The presence of ultradian, circadian and infradian frequency band oscillations were identified using Thomson F-test multi-taper scheme^48,49^. (**a**) Preprocessed single impedance signal (z-score and down sampled to 48 samples/day). A 5-day moving window (moving step of 5 days) was used to identify significant cycles in ultradian and circadian frequency bands. A 100-day moving window (moving step of 5 days) was used for cycles in infradian band. (**b**) Periodicity test in ultradian band for each 5-day signal segment was investigated using Thomson F-test based on the multi-taper scheme^48,49^ to test the periodicity at the frequencies of interest (FOI, see below). The time-half-band product (TW) was set at three (TW = 3) and the number of tapers (K) was five (K = 5), resulting in the frequency resolution of 2W = 1.2 cycles/day. In ultradian band, FOI was chosen uniformly spaced in periods, from 23/24 = 0.96 hours/cycle to 6 hours/cycle with an incremental step of 15 minutes. At each fixed frequency, the F-test was set at the level of upper 99% (*p* = 0.01) percentage point of the F-distribution with 2 and 2K-2 = 8 degrees of freedom under the null hypothesis of no spectral line, leading to the critical value = 8.02. The upper panel shows the F-statistic in each 5-day window throughout the duration of the signal, where the red dash indicates the F-statistic above the critical value (see scale color bar). The series blue dots below indicate the time instance of ultradian periodicity detected. A dot means at least one of the FOIs has shown reliable periodicity in the 5-day window. **(c)** Periodicity test at circadian frequency. The green curve shows the F-statistic at the frequency of 1 cycle/day and the red horizontal line is the critical value of F-statistic, which is the same as that for ultradian band. The series of red dots below indicates the time instances when the F-statistic was above the critical value. Note that due to the uncertainty of estimated frequency quantified by the half-band width W = 3/5 = 0.6 cycles/day, a positive test indicates the frequency range of detected periodicity is in 1 ± 0.6 cycles/day. (**d**) Periodicity test in infradian band. Same procedure as for the tests in ultradian band, except that the window length was 100 days and FOI was from 2.5 days/cycle to 30 days/cycle with an incremental step of 15 minutes. In the upper panel, the red dots indicate the F-statistic was above the critical value, same as those for ultradian and infradian testing, while the series of brown dots indicate whether a periodicity shown in any of the FOIs in the infradian band.

## Acknowledgements

We deeply appreciate the efforts of our patients who made this research possible. We thank 1) Mayo Clinic research team and in particular the efforts of Karla Crockett, Cindy Nelson, and Starr Guzman. 2) The Medtronic Inc. team for providing the investigational Medtronic Summit RC+S™ devices and engineering support from Abbey Becker, PhD and Dave Linde, PhD. The research was primarily funded by NIH Brain Initiative UH2&3 NS095495, R01 NS112144, and R01 NS092882. Additional funding for analysis of impedance was provided by DARPA HR0011-20-2-0028), Mayo Clinic philanthropy, and community expertise and resources made available by the NIH Open Mind Consortium NIH U24-NS113637 (https://openmindconsortium.github.io/). V.K. was partially supported by the Czech Technical University, Prague, Czech Republic.

## Author contributions

Conceptualization, project administration and supervision:

GAW, VK, MS, and TD

Methodology:

Investigation (data collection): FM, VK, VS, JC, NG, IB, TM, PEC, BNL, DH, SM, BHB, KJM, JVG, MS, GAW.

Data curation: FM, VK, VS, TM, BNL, DH, SM, BHB, TD, KJM, JVG, MS, GAW.

Formal analysis: FM, VK, VS, JC, GAW.

Validation: FM, VK, VS, JC, GAW.

Visualization: FM, VK, VS, BHB

Software: FM, VK, VS, JC. BHB, DH

Writing— original draft: FM, VK, GAW.

Writing—review and editing: FM, VK, VS, JC, NG, IB, TM, EKS, PEC, BNL, NN, DH, SM, SW, TR, BHB, TD, KJM, JVG, MS, GAW.

